# The ratio of left atrial and ventricular volume as new marker of atrial cardiopathy with relevance for stroke risk and cognitive dysfunction

**DOI:** 10.64898/2025.12.18.25342359

**Authors:** Julian Deseoe, Martin Hänsel, Lisa Herzog, Neda Davoudi, Andreas R. Luft, Catherine Gebhard, Bjoern Menze, Beate Sick, Susanne Wegener

**Affiliations:** Department of Neurology and Clinical Neuroscience Center, University Hospital Zurich and University of Zurich, Switzerland; Neuroscience Center Zurich, University of Zurich, Zurich, Switzerland; ETH Zurich, Institute for Biomedical Engineering, Department of Information Technology and Electrical Engineering, Zurich, Switzerland; Department of Quantitative Biomedicine, University of Zurich, Zurich, Switzerland; cereneo Center for Neurology and Rehabilitation, Vitznau, Switzerland; Department of Cardiology, Inselspital University Hospital Bern, University of Bern, Bern Switzerland; Epidemiology, Biostatistics & Prevention Institute, University of Zürich, Zürich, Switzerland; TIDIT, Thurgau Institute for Digital Transformation, Kreuzlingen, Switzerland

## Abstract

**Background:** Atrial cardiopathy is an important cause of embolic stroke and a potential cause of cognitive dysfunction. Increased left atrial volume indexed to body surface area (LAVi) has been widely used as marker for atrial cardiopathy. However, since physiological remodeling may also increase LAVi, it lacks specificity. Left atrial to left ventricular volume (LA:LV) ratio was recently suggested as a better marker of atrial cardiopathy allowing a more accurate detection of imbalanced, pathological atrial remodeling. We investigated if LA:LV ratio is associated with different established sequelae of atrial cardiopathy and if it is superior to LAVi for the detection of atrial fibrillation/flutter (AF) as cause of stroke.

**Methods:** We compared the association of LAVi and LA:LV ratio with risk of ischemic stroke or transient ischemic attack (TIA) in the population-based UK Biobank cohort (n= 38’848), using cause specific hazard models. In addition, we tested for an association with cognitive function using multivariate linear regression models. Finally, we investigated the association of LAVi and LA:LV ratio with probability of atrial fibrillation/flutter being identified as stroke etiology in a cohort of ischemic stroke patients (n = 1’273). We compared the test performance of LAVi and LA:LV ratio for identifying atrial fibrillation/flutter as stroke etiology.

**Results:** While LAVi was not significantly associated with risk of ischemic stroke/TIA (HR: 1.11, 95% CI 0.97 - 1.26, p = 0.14), LA:LV ratio was (HR: 1.15, 95% CI 1.01-1.30, p = 0.04). Besides, LA:LV ratio was more strongly associated with worse cognitive function in healthy adults [LA:LV ratio: -0.024 (95% CI (- 0.032) - (- 0.015)), LAVi: - 0.010 (95% CI (- 0.019) - (- 0.002))]. In a stroke patient cohort, LA:LV ratio showed a significantly better performance in identifying AF as underlying cause of ischemic stroke compared to LAVi (Receiver Operating Characteristic Area under the Curve 0.856 (0.803- 0.908) vs. 0.808 (0.750 - 0.866) p = 0.03).

**Conclusions:** We provide evidence that LA:LV ratio is a promising marker of atrial cardiopathy. Hence, LA:LV ratio has the potential to improve diagnosis of atrial cardiopathy, facilitating prevention of ischemic stroke and maintaining of brain health.

## 1. Introduction

Atrial cardiopathy is a structural and functional disorder of the atria, which is seen as both potential cause and consequence of atrial fibrillation (AF).^1^ It has also been associated with higher risk of stroke^2^ as well as lower cognitive performance,^3^ even in the absence of AF. One leading explanatory hypothesis is that due to remodeling in atrial cardiopathy, thrombogenicity of the atria increases, leading to increased risk of stroke.^4^ Similarly, cerebral micro-embolism due to atrial cardiopathy may lead to cognitive decline even in the absence of ischemic stroke.^3,5,6^ Atrial cardiopathy has also been linked to cortical atrophy.^3^ There is evidence that anticoagulation may reduce future ischemic strokes in patients with atrial cardiopathy, even in the absence of atrial fibrillation or flutter (AF).^7^ However, the diagnosis of atrial cardiopathy in the absence of AF is not yet reliable enough to justify anticoagulation.

Although the importance of atrial cardiopathy is increasingly apparent, reliable methods for its diagnosis are still sparse.^4^ Large left atrial volume indexed to body surface area (LAVi) has been commonly used as an indicator of atrial cardiopathy.^8^ Increased LAVi has been associated with increased risk of ischemic stroke^9-11^ and poorer cognitive function,^3,12,13^ although there has been variation between studies. In clinical practice, increased LAVi can indicate ischemic stroke patients with high likelihood of AF as underlying stroke cause.^14^

However, the use of LAVi for the detection of atrial cardiopathy has important limitations. Physiological remodeling of the left atrium, for example due to physical activity, can also lead to increased LAVi complicating its use as a marker of pathological remodeling.^15-19^ Recently, the left atrial to ventricular volume (LA:LV) ratio, defined as the ratio between the maximal left atrial volume at end systole and left ventricular end diastolic volume (LVEDV) has been proposed to better differentiate between balanced, physiological and imbalanced, pathological left atrial enlargement.^19-21^ This is based on the observation that physiological remodeling, such as that induced by exercise, typically results in proportional increases in both left atrial volume and left ventricular volume, thereby maintaining a relatively stable LA:LV rato.^15-17,19^ However, associations of LA:LV ratio with known sequelae of atrial cardiopathy impacting brain health have never been systematically studied.

We aimed to compare LA:LV ratio and LAVi as markers of atrial cardiopathy, by investigating their associations with different sequelae of atrial cardiopathy. First, we evaluated the associations of LAVi and LA:LV ratio with the risk of ischemic stroke or transient ischemic attack (TIA), cognitive function and cortical atrophy in subpopulations of the population-based UK Biobank (UKBB) cohort. Second, we examined the predictive performance of LAVi and LA:LV ratio for identifying AF as the underlying cause of stroke in a clinical cohort of ischemic stroke patients from the University Hospital Zurich (USZ).

## 2. Methods

### Association between left atrial indices and risk of ischemic stroke/TIA, cognitive function and cortical atrophy in the UKBB

The UKBB is a large population-based cohort study, which holds data of 502’128 participants from the UK. The recruitment protocol is publicly available.^22^ The UKBB imaging study contains among other assessments, cardiac magnetic resonance (CMR) imaging for over 50’000 participants.^23^ We worked with the data release from March 2025 (v19). From the CMR imaging cardiac chamber volumes have been previously extracted using deep learning and made accessible to researchers.^24^ LAVi and LA:LV ratio were standardized to have a mean of zero and a standard deviation of one to facilitate comparison of effect sizes.

### Participant selection

Data of all participants with available information on LA volume were considered for analysis. For assessment of associations with risk of stroke/TIA, patients with AF diagnosed before the imaging date, as well as patients with heart failure were excluded. Participants with AF were excluded as quantification of cardiac volumes with CMR in these patients can be imprecise.^25^ Patients with heart failure were excluded as the interplay between left atrial volume and left ventricular volume changes in these patients.^26^ Hence, these likely warrant a separate investigation. In addition, participants with prior stroke or TIA were excluded. All participants with incomplete data were excluded. Details of how participants with AF and heart failure were identified can be found in the supplemental methods.

For assessment of association with cognitive function, the same inclusion and exclusion criteria were applied with two exceptions. Patients with prior TIA were not excluded while those with preexisting dementia were excluded. An overview of inclusion and exclusion of participants can be found in Figure 1.

**Figure 1.**
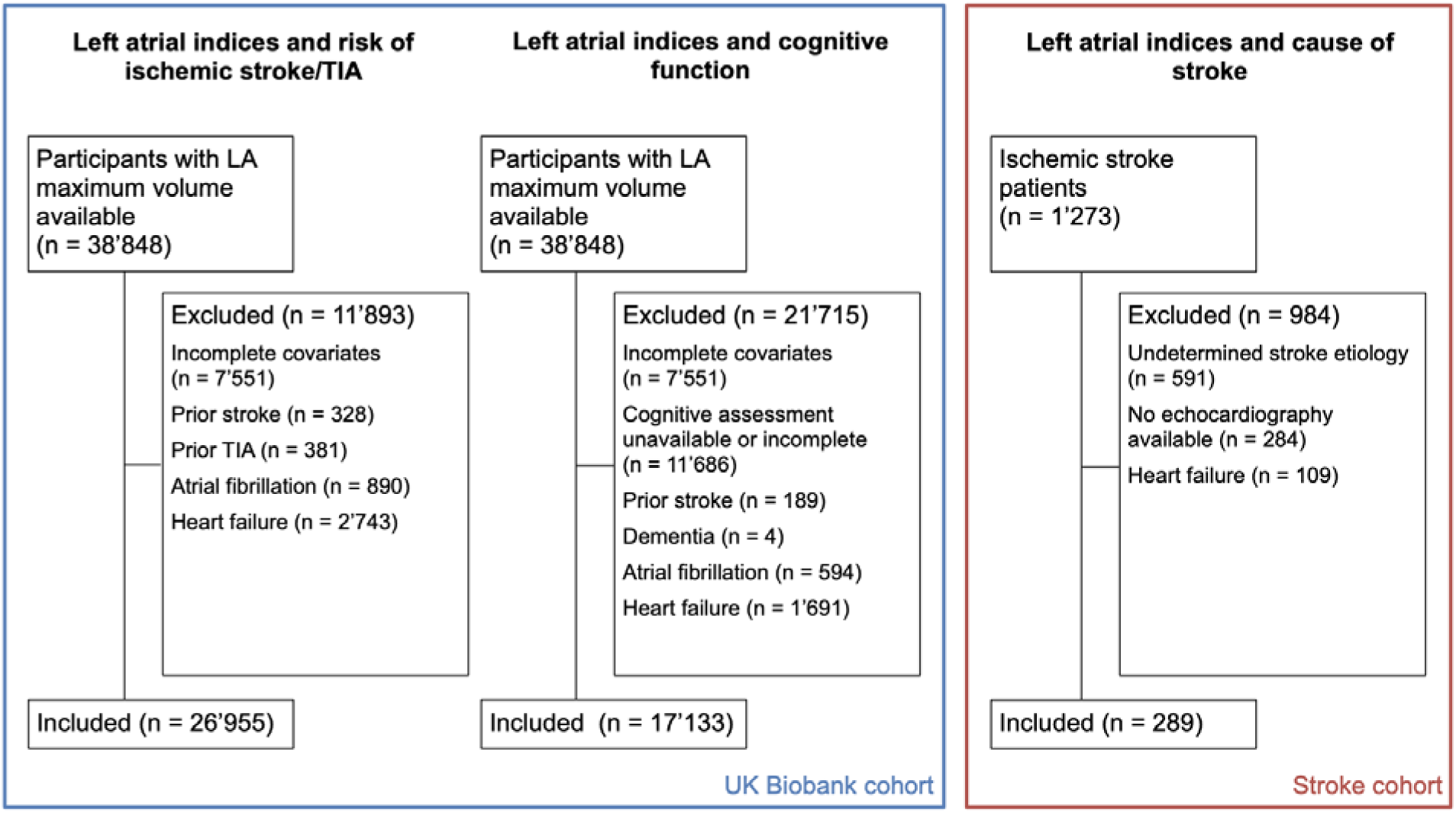
Inclusion flow chart. LA: left atrium

### Statistical Analysis

All analyses were performed using R Studio with R version 4.4.0. Details on the definition of endpoints can be found in the supplemental material. For assessment of associations of LAVi and LA:LV with ischemic stroke/TIA risk, a competing risks analysis treating ischemic stroke/TIA and death as competing events was performed. We calculated cause specific and subdistribution hazard models^27^ and performed the same regressions for the competing event, death. We employed both unadjusted models and models adjusted for the covariates shown in Table 1.

**Table 1.**
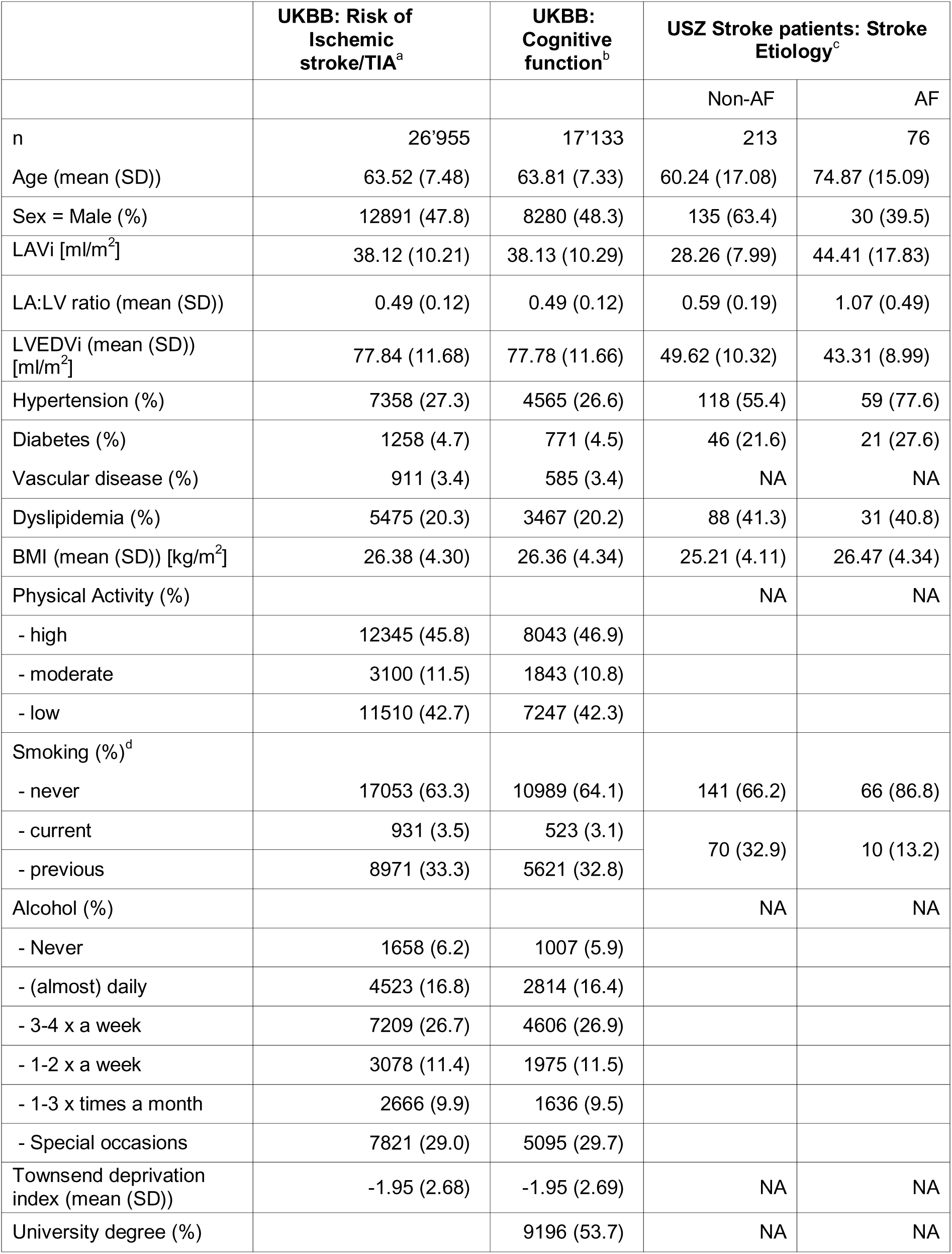
Baseline Characteristics of the studied cohorts. UKBB: UK Biobank, AF: Atrial fibrillation/flutter, LAVi: left atrial volume indexed to body surface area, LVEDVi: left ventricular end-diastolic volume indexed to body surface area, LA:LV ratio: left atrial to ventricular volume ratio. SD: standard deviation ^a^ “UKBB: Ischemic stroke” is a subpopulation of the UK Biobank cohort used for analysis of association of ischemic stroke risk and atrial indices. ^b^ “UKBB: Cognitive function” is a subpopulation of the UK Biobank used do assess association of atrial indices and cognitive function. ^c^ “USZ stroke patients: Stroke etiology” is a cohort of ischemic stroke patients from the USZ. ^d^ For ischemic stroke patients, smoking status was only available classified as current/previous or never. For two patients smoking status was not available.

Details on ascertainment of covariates can be found in the supplemental material and Table S1. We tested for violation of the proportional hazard assumption by assessing Schoenfeld residuals.^28^ We investigated sex differences by including an interaction term between LA:LV ratio and sex and LAVi and sex respectively in separate models.

Linear regression models were calculated to assess the association of LAVi and LA:LV ratio with cognitive function, both in unadjusted and fully adjusted models. We investigated a potential nonlinear association between LAVi or LA:LV ratio and cognitive function using restricted cubic spline with three nodes.^29^ Significance of nonlinear components was assessed using partial F test. Finally, we investigated whether there was an interaction between sex and LAVi or sex and LA:LV ratio for the association with cognitive function.

We then assessed the associations of LAVi and LA:LV ratio with magnetic resonance image measured volumes of 66 cortical parcellations according to Desikan-Killiany parcellation^30^ using linear regression models adjusted for age, sex and intracranial volume. We adjusted for multiple testing using the Benjamini-Hochberg procedure.^31^ The temporal pole parcellation is not available in the UKBB data due to its unreliable segmentation and was thus excluded. Details can be found in the Supplemental Methods. We again excluded all participants with previous stroke, AF, heart failure or missing data.

### Left atrial indices for detection of AF as cause of stroke in ischemic stroke patients from the USZ

Data of all patients who had not refused consent for use of their routine data for research, treated for ischemic stroke at the USZ in the years 2023 and 2024 were retrospectively collected. We received permission from the Cantonal Ethics committee Zurich to perform this data collection (PREDICT, BASEC-ID: PB_2016_01751). Cardiac volumes were extracted from echocardiography reports, routinely performed after ischemic stroke. For the values to be included in the study, the echocardiography had to be performed after the ischemic stroke diagnosis and before the outpatient checkup at approximately three months after stroke. LAV was measured using the biplane method, at the end of systole.^32,33^ Patients with heart failure were once again excluded. Details can be found in the supplemental methods.

Stroke etiology was identified from three-month checkup report if available or discharge report. Similar to previous studies,^14^ stroke etiology was categorized into three subgroups: 1) AF, 2) other determined cause and 3) unknown etiology/incomplete workup. Detailed criteria are shown in Table S2. Patients with unknown etiology or incomplete work up were excluded from the analysis. An overview can be found in Figure 1.

Prior diseases reported in Table 2, BMI, age and sex were extracted from the Swiss Stroke registry.

**Table 2.**
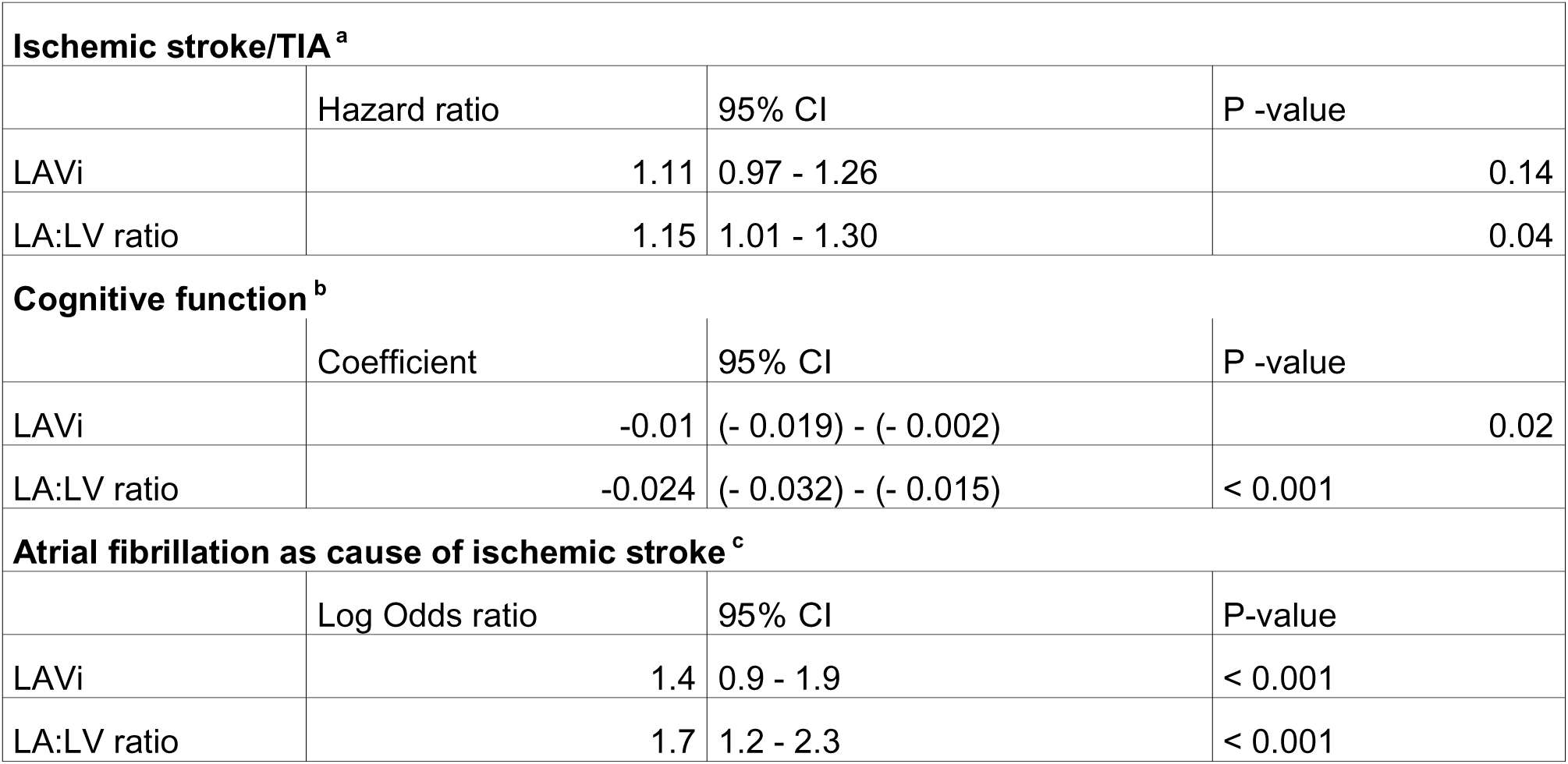
Associations of standardized left atrial volume index and standardized left atrial to left ventricular volume ratio with sequelae of atrial cardiopathy. LAVi: left atrial volume index, LA:LV ratio: left atrial to ventricular volume ratio ^a^ Hazard ratios from cause specific hazard models for ischemic stroke with death as competing event. HR > 1 indicate increased hazard rate of ischemic stroke/TIA. For LAVi and LA:LV ratio two separate models were calculated each adjusted for the described covariates. 249 participants had an ischemic stroke/TIA as an endpoint (TIA: 88, ischemic stroke: 161), 606 deaths. Median time under observation was 6.8 years. ^b^ Linear regression coefficients for cognitive function as independent variable. Coefficients > 0 indicate association of larger volumes with better cognitive function. For LAVi and LA:LV ratio two separate models were calculated each adjusted for the described covariates. ^c^ Log odds ratio for AF as stroke cause as independent variable in multivariate logistic regression. Coefficients > 0 indicate association of larger volumes with AF as stoke cause. For LAVi and LA:LV ratio two separate models were calculated each adjusted for age and sex. 289 ischemic stroke patients included, 76 with atrial fibrillation as stroke cause.

### Statistical analysis

Logistic regression models for the association of standardized LAVi and LA:LV ratio with AF being identified as stroke cause were calculated for LAVi and LA:LV ratio, both in unadjusted models and adjusting for age and sex. Area under the receiver operating characteristic curves (ROC-AUC) were calculated for LAVi and LA:LV ratio for classifying stroke cause as either AF or other determined stroke etiology and compared using DeLongs test.^34^ Then, the ideal cut off point in our sample for both parameters was calculated by maximizing the Youden index.^35^ Using 10- fold cross validation, sensitivity and specificity as well as positive and negative likelihood ratios were calculated for the two parameters. We also evaluated predictive performance in male and female patients separately.

Finally, we investigated if LA:LV ratio held complimentary information to MR proANP, a blood biomarker, which is also used to indicate atrial cardiopathy.^36^ Details can be found in the supplemental material.

## 3. Results

Figure 1 shows the inclusion/exclusion flow chart for participants included into this study. For the analysis of atrial indices and risk of ischemic stroke/TIA, 26’955 participants were included. For analysis of cognitive function, 17’133 participants were included. Although both analyses started with the same initial UKBB dataset, a substantial number of participants lacked complete cognitive assessments, resulting in a smaller final sample size. A summary of cohort characteristics is shown in Table 1.

Cardiac chamber volumes as measured by echocardiography in the clinical cohort were smaller compared to the UKBB cohort (LAVi: 38 ml/m^2^ vs 32 ml/m^2^, LVEDVi: 78 ml/m^2^ vs 48 ml/m^2^), which is consistent with the known tendency of echocardiography, to underestimate chamber volumes compared to CMR.^37^

We visually investigated the association of LAVi and LA:LV ratio with age, sex and everyday physical activity (Figure 2). LAVi showed a positive association with physical activity. Women had larger mean LA:LV ratios than men, while no such sex difference was observed for LAVi. LAVi has been shown to be similar between sexes while men typically have larger LVEDVi, explaining our findings.^38^ These results were also confirmed using linear regression models (Tables S3-S4). To account for potential sex differences, all subsequent analysis were controlled for sex and investigated for sex specific effects. So cardiac volumes reported in this study confirmed findings from existing literature.

**Figure 2.**
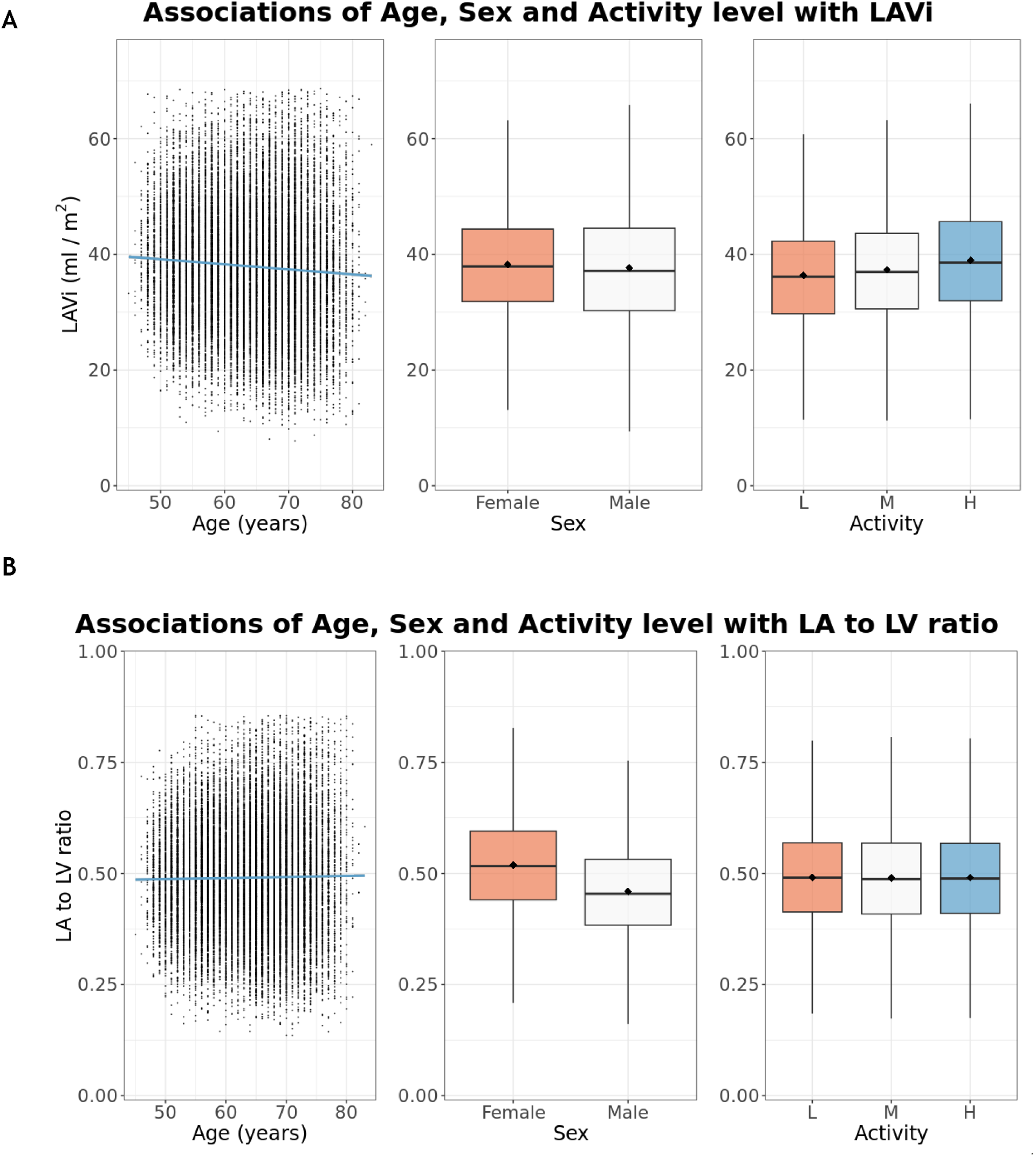
(A) Association of Age, Sex and Activity with left atrial volume indexed to body surface area. (B) Association of Age, Sex and Activity with left atrial to ventricular volume ratio. Results from UK Biobank cohort used for ischemic stroke/TIA analysis. Blue lines show linear regression lines. For LAVi and LA:LV ratio outliers more than three standard deviations from the mean are not shown. Boxplots show mean (point) and median (line) values with interquartile range. Values outside two standard deviations from the mean are not shown. LAVi: left atrial volume indexed to body surface area, LA to LV ratio: left atrial to ventricular volume ratio, L: low, M: medium, H: high.

We then investigated the association of LA:LV ratio and LAVi with risk of ischemic stroke/TIA. In the UKBB cohort, LAVi was not significantly associated with incident ischemic stroke both in unadjusted (Table S5) and fully adjusted models (Table 3). In contrast, larger LA:LV ratio was significantly associated with an increased risk of future ischemic stroke/TIA (aHR 1.15, 1.01-1.30, p = 0.04). When additionally including interaction terms with sex to the adjusted models, we observed no significant interactions between sex and LAVi (p = 0.14) and sex and LA:LV ratio (p = 0.83), meaning that there is no significant evidence for sex differences for these associations. No significant associations were found between cardiac volumes and mortality (Table S6). Results for subdistribution hazard models showed similar results (Tables S7-S8).

**Table 3.** Test metrics of LA:LV ratio and LAVi for detecting atrial fibrillation as stroke cause with 95% CI. LA:LV ratio: left atrial to ventricular volume ratio, LAVi: left atrial volume indexed to body surface area, LR +: positive likelihood ratio, LR –: negative likelihood ratio ^a^ Optimal cut o□ calculated by maximizing Youden Index 95% CI calculated using 1000 bootstrap samples. ^b^ Test metrics calculated using 10-fold cross validation. 95% CI are Wilson CI.

|  | LA:LV ratio | LAVi |
| --- | --- | --- |
| Optimal cut off point <sup>a</sup> | 0.81 (0.68 - 0.87) | 35 ml/m<sup>2</sup> (34 -36) |
| Sensitivity <sup>b</sup> | 0.684 (0.573 -0.778) | 0.658 (0.546 - 0.755) |
| Specificity <sup>b</sup> | 0.850 (0.796 - 0.892) | 0.808 (0.749 - 0.855) |
| LR + <sup>b</sup> | 4.6 (3.3 - 6.7) | 3.4 (2.5 - 4.8) |
| LR - <sup>b</sup> | 0.37 (0.25 - 0.50) | 0.42 (0.29 - 0.56) |

Atrial cardiopathy was previously found to be associated with cognitive decline and cortical atrophy, potentially caused by cerebral microembolism.^3^ Therefore, we tested for associations of LAVi and LA:LV ratio with cognitive function. Larger LA:LV ratio was more strongly associated with poorer cognitive function [Regression coefficient: - 0.024, 95% CI (- 0.032) - (- 0.015)] than LAVi [Regression coefficient: - 0.010, 95%CI (- 0.019) - (- 0.002)] in fully adjusted (Table 3) and unadjusted models (Table S5). No significant interactions for sex and LAVi (p = 0.83) or LA:LV ratio (p = 0.70) were observed. We found no evidence of nonlinearity. (Figure S1).

Larger LA:LV ratio was associated with cortical atrophy especially in the temporal lobe, insula and cingulate cortex. Associations with LAVi were partially present, but weaker (Figure 3, Supplemental Data). Interestingly, atrophy in temporal lobe, insula and cingulate cortex have been previously linked to AF and atrial cardiopathy,^39-40^ so that the cortex-specific associations found with LA:LV ratio could potentially represent an atrial cardiopathy related brain imaging phenotype.

**Figure 3.**
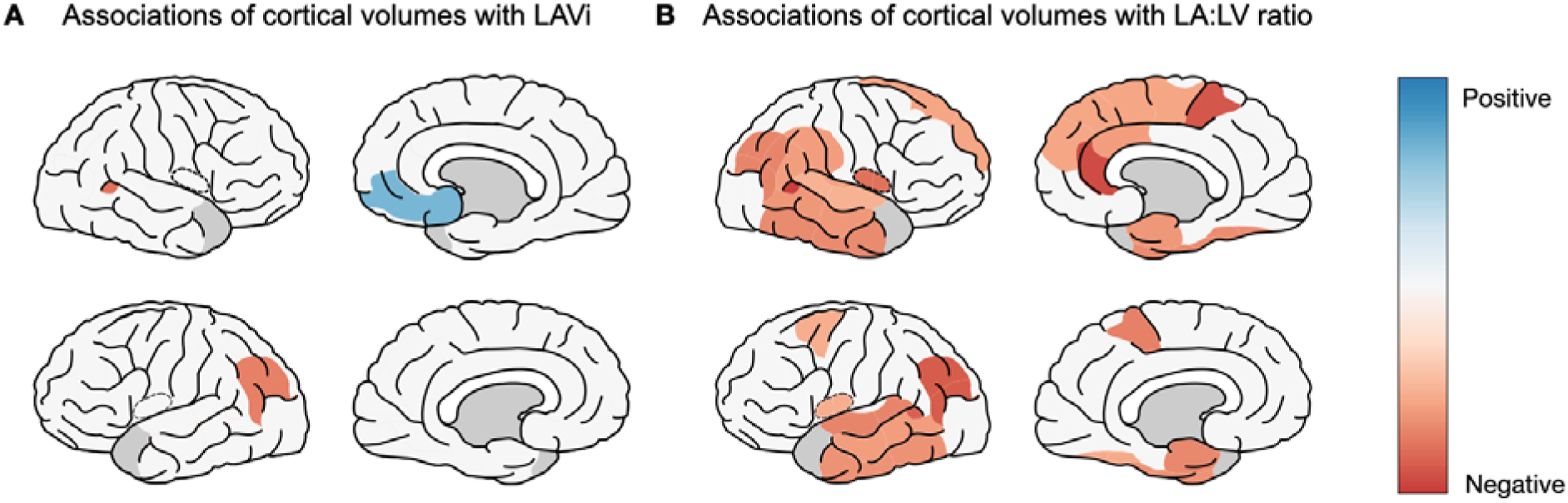
(A) Associations of LAVi with cortical volumes according to Desikan-Killiany parcellation. (B) Associations of LA:LV ratio with cortical volumes according to Desikan-Killiany parcellation. Colors show significant (p < 0.05) regression coefficients for association of standardized cortical volume with standardized LAVi/LA:LV ratio, adjusted for age, sex and intracranial volume. Red regions show atrophy associated with larger LAVi/LA:LV ratio. Parcellation of the temporal pole (grey) was not performed in the UK Biobank data due to unreliable results. Analysis included 28’954 participants with available heart and brain imaging. Plot created using python package pySimpleBrainPlot. LAVi: left atrial volume indexed to body surface area, LA to LV ratio: left atrial to ventricular volume ratio.

Next, we examined the association of standardized LAVi and LA:LV ratio with the probability of AF being identified as the cause of stroke in a cohort of 289 ischemic stroke patients from the USZ. Results are shown in Table 3. Both standardized LAVi and LA:LV ratio were strongly associated with AF as the stroke cause, with the effect size of LA:LV ratio being slightly larger in adjusted modes (LA:LV ratio: Log Odds ratio: 1.7, 95% CI 1.1 – 2.3, p < 0.001, LAVi: Log Odds ratio: 1.4 95% CI 0.9 - 1.9, p < 0.001) and unadjusted models (Table S4). The LA:LV ratio showed a significantly higher ROC-AUC compared to the LAVi for identifying AF as the stroke cause (0.856 (0.803 - 0.908) vs. 0.808 (0.750 - 0.866) p = 0.03) (Figure 4). LA:LV ratio outperformed LAVi on all calculated test metrics (Table 3).

**Figure 4.**
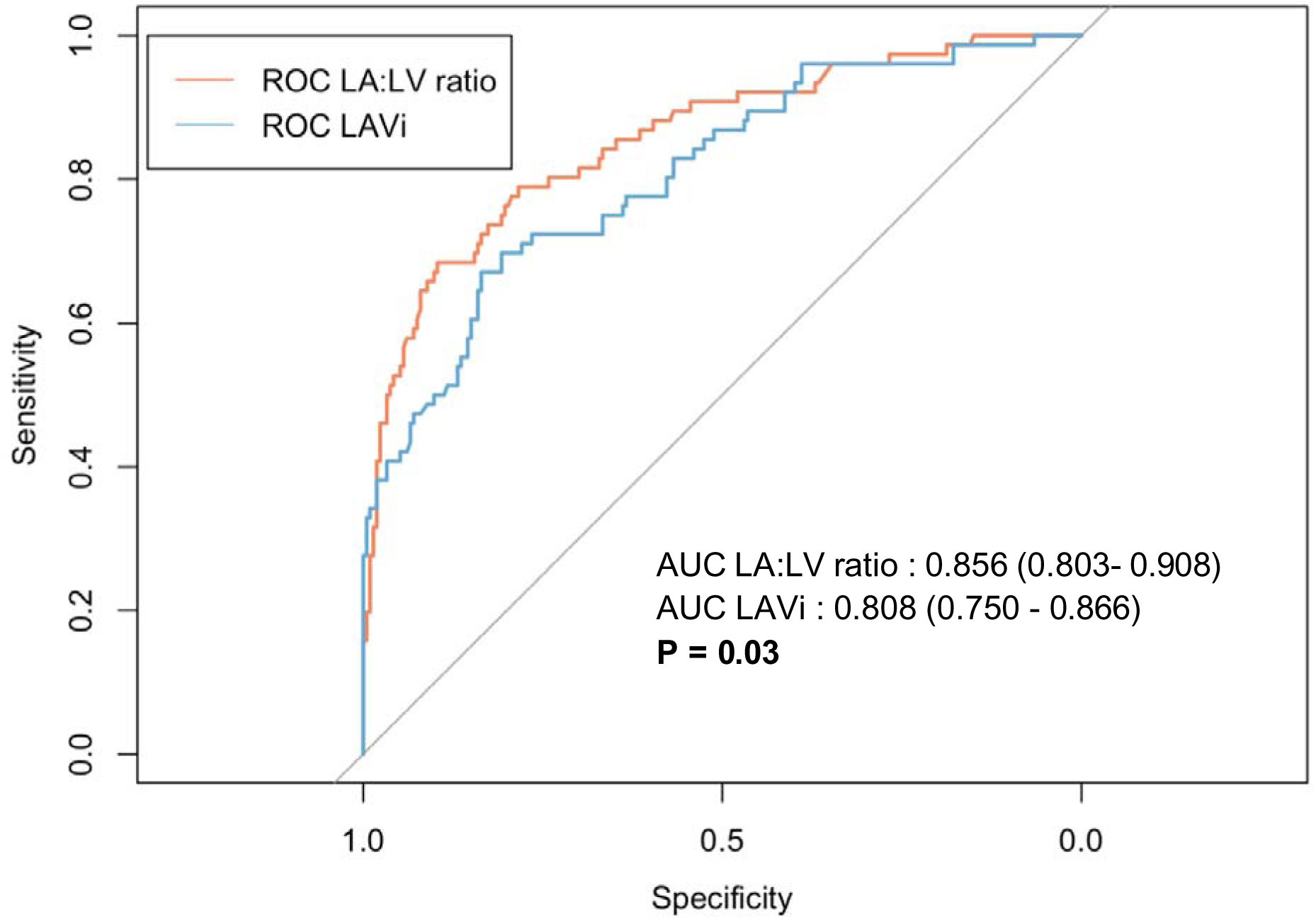
Receiver operating characteristic curves for LAVi and LA:LV ratio for detecting atrial fibrillation as stroke cause. Area under the curve with 95 % CI is shown in the bottom right. Confidence intervals were calculated based on 1000 bootstrap samples. P value calculated with DeLong’s test. LAVi: Left atrial volume indexed to body surface area. LA:LV ratio: Left atrial to ventricular volume ratio, ROC: Receiver operating characteristic, AUC: Area under the Curve.

A sex stratified analysis showed that LA:LV ratio showed higher ROC-AUC for identifying AF as the stroke cause than LAVi in both sexes. The ideal cut-off as calculated by the Youden Index was higher for women compared to men (Women: 0.85 95% CI 0.69 – 0.96, Men: 0.71 95% CI 0.55 – 0.86), (Table S9), which aligns with findings from the UKBB, were women showed higher average LA:LV ratio.

Finally, we aimed to capture the effect of combining LA-LV with current clinical and blood biomarkers for prediction of atrial cardiopathy. In a logistic regression model including LA:LV ratio, MR proANP, age and sex, both higher MR proANP and higher LA:LV ratio were significantly associated with an increased likelihood of AF as stroke cause. (Table S10) The multivariate model showed an improved ROC-AUC for predicting AF as stroke cause (0.897, 95% CI 0.849 - 0.945) compared to LA:LV ratio alone although the difference was not statistically significant. (p = 0.28). (Figure S2).

## 4. Discussion

We found that in different cohorts and independent of assessment modality (echocardiography versus CMR), the LA:LV ratio consistently showed stronger associations than LAVi with clinically relevant manifestations of atrial cardiopathy. In the UKBB, higher LA:LV ratios were linked to greater risk of ischemic stroke/TIA and poorer cognitive function. Further, larger LA:LV ratio was significantly associated with cortical atrophy especially in temporal lobe, insula and cingulate cortex, providing a potential link between larger LA:LV ratio and worse cognitive function. In stroke patients, the LA:LV ratio showed improved diagnostic performance for detecting AF as the stroke cause, with higher ROC-AUCs in both sexes and sex-specific optimal cut-offs.

Our findings align with our hypothesis that LA:LV ratio is more strongly associated with sequelae of atrial cardiopathy than LAVi and show its potential as a marker for atrial cardiopathy. We hypothesize that this is because it addresses key limitations of LAVi for assessing atrial cardiopathy as it allows improved detection of imbalanced, pathological cardiac remodeling There is abundant evidence that left atrial, and the left ventricular function are interdependent.^41^ For example, changes in left atrial function influence left ventricular filling pressure and vice versa.^41.42^ The complex interplay of left atrium and ventricle is not captured by measuring LAVi alone. In contrast, while no single marker could ever fully characterize these interactions, the LA:LV ratio is a straightforward way of assessing the left atrium, while incorporating information from the left ventricle.

We hypothesize that, as it allows joint assessment the left atrium and ventricle, the LA:LV ratio can better distinguish between balanced and unbalanced cardiac remodeling. While there is little literature directly comparing the effects of balanced and unbalanced atrial remodeling on the risk of cardiovascular events, there is some evidence comparing the effects of left atrial enlargement (LAE) in physically active and inactive people. Participants with LAE who were physically active, had a significantly lower risk of AF compared to inactive participants with LAE.^43^ Further, physical activity has been associated with reduced risk of AF and stroke.^44,45^ Therefore, an increased LAVi with unchanged or decreased LVEDVi may better represent an atrial cardiopathy phenotype, which would be associated with increased risk of stroke, worse cognitive function and increased probability of AF as stroke cause. Large LAVi coupled with increased LVEDVi, on the other hand, may be more representative of physiological changes, e.g. induced by increased physical activity, and thus less indicative of adverse cardiovascular events. However, more, especially longitudinal studies are needed to further investigate this hypothesis.

The LA:LV ratio could easily be incorporated into clinical practice, since the required measures LAVi and LVEDVi are routinely obtained in echocardiography or cardiac imaging. We demonstrate this by validating LA:LV ratio in a clinical cohort. In patients with ischemic stroke, early detection of atrial cardiopathy is essential, as patients with atrial fibrillation require anticoagulation to be adequately protected from stroke recurrence.^46^ LA:LV ratio could improve selection of patients for long term rhythm monitoring to detect underlying AF. Increased LA:LV ratio might even become a sufficient justification for prescribing anticoagulation in ischemic stroke patients, especially in patients, where no other convincing stroke etiology is found, as atrial cardiopathy can lead to cardioembolic stroke even in the absence of AF.^7^ Hence, this study should inspire prospective multicenter trials to facilitate the establishment of LA:LV ratio in clinical practice, especially as part of diagnostic work-up of ischemic stroke.

### Limitations

Due to the large sample size of the UKBB, statistically significant but not clinically relevant associations might be detected. This is especially a potential limitation for associations of LA:LV ratio with ischemic stroke/TIA in the UKBB cohort, where effect size was only borderline significant. We addressed this by also evaluating LAVi and LA:LV ratio in a smaller clinical cohort. Another limitation of our study is that we deliberately excluded subjects with heart failure, reduced EF and LV dilation, as interaction between LAVi and LVEDVi are different in these patients. These subjects should be studied in dedicated cohorts. Finally, although longitudinal studies have been conducted on effects of exercise on heart volumes,^15-17,19^ supporting our mechanistic hypothesis, establishing causality is not possible with an observational, cross-sectional approach. Further, it would have been interesting to correlate and compare LA:LV ratio with other proposed imaging markers of atrial cardiopathy such as left atrial strain.^47^ However, left atrial strain was not measured in the UKBB imaging protocol and is not routinely measured in clinical practice at the University Hospital Zurich.

### Conclusions

We show that LA:LV is a promising new marker of atrial cardiopathy and demonstrate its associations with clinically relevant manifestations of atrial cardiopathy. Further, we show its potential value for guiding diagnostic workup post ischemic stroke. Prospective, multicenter studies are needed to further validate our findings and establish sex specific reference values to facilitate the use of LA:LV ratio in clinical practice.

## 5. Acknowledgements

This research has been conducted using the UK Biobank Resource under application number 467527. This work uses data provided by patients and collected by the NHS as part of their care and support. The clinical data analyzed in this study will be made available upon reasonable request. Figure layout was created using Biorender

## 6. Data Availability Statement

The UK Biobank will make the data available to all bona fide researchers for all types of health-related research that is in the public interest. For more details on the access procedure, see the UK Biobank website: http://www.ukbiobank.ac.uk/register-apply/. Anonymized data from the University Hospital Zurich not published within this article will be made available by request from any qualified investigator.

## 6. Sources of Funding

This work was supported by the UK Biobank Platform Credits Programme for covering computational costs, and the Swiss National Science Foundation (SNSF 310030_200703) and the UZH Candoc Grant (25-024)

## 7. Disclosures

None

## 8. Supplemental Material

Detailed Methods

Tables S1-S10

Figures S1-S2

Supplemental Data

## Notes

### Competing Interest Statement

The authors have declared no competing interest.

### Author Declarations

The Cantonal Ethics committee Zurich gave ethical approval for this work. Further the UK Biobank data Access committee gave its approval (Project ID 467527)

### Summary of Updates

Added additional considerations to the Discussion; Methods updated to improve clarity; Author affiliations updated; Supplemental filed updated.

