## Supplemental Data for "The ratio of left atrial and ventricular volume as new marker of atrial cardiopathy with relevance for stroke risk and cognitive dysfunction"

| | Left atrial volume index | | | Left atrial to ventricular volume ratio | | |
|---|---|---|---|---|---|---|
| Parcellation | Coefficient | P-values | Adjusted P-values | Coefficient | P-values | Adjusted P-values |
| bankssts (left) | -0.013 | 0.015 | 0.142 | -0.021 | < 0.001 | **0.001** |
| caudalanteriorcingulate (left) | 0.001 | 0.893 | 0.936 | -0.004 | 0.455 | 0.546 |
| caudalmiddlefrontal (left) | -0.011 | 0.033 | 0.231 | -0.013 | 0.011 | **0.032** |
| cuneus (left) | 0.009 | 0.106 | 0.306 | 0.007 | 0.188 | 0.354 |
| entorhinal (left) | -0.002 | 0.746 | 0.879 | -0.017 | 0.003 | **0.013** |
| fusiform (left) | -0.002 | 0.636 | 0.824 | -0.014 | 0.005 | **0.018** |
| inferiorparietal (left) | -0.018 | 0.001 | **0.013** | -0.022 | < 0.001 | **< 0.001** |
| inferiortemporal (left) | -0.007 | 0.140 | 0.356 | -0.016 | 0.001 | **0.004** |
| isthmuscingulate (left) | 0.003 | 0.530 | 0.769 | 0.005 | 0.359 | 0.490 |
| lateraloccipital (left) | -0.001 | 0.798 | 0.924 | -0.001 | 0.805 | 0.839 |
| lateralorbitofrontal (left) | -0.001 | 0.855 | 0.931 | -0.008 | 0.084 | 0.181 |
| lingual (left) | 0.010 | 0.074 | 0.257 | 0.002 | 0.772 | 0.835 |
| medialorbitofrontal (left) | 0.012 | 0.015 | 0.142 | -0.003 | 0.561 | 0.638 |
| middletemporal (left) | -0.009 | 0.065 | 0.252 | -0.015 | 0.002 | **0.007** |
| parahippocampal (left) | 0.005 | 0.414 | 0.739 | -0.006 | 0.336 | 0.472 |
| paracentral (left) | -0.004 | 0.453 | 0.743 | -0.018 | 0.001 | **0.004** |
| parsopercularis (left) | -0.004 | 0.501 | 0.767 | -0.003 | 0.640 | 0.716 |
| parsorbitalis (left) | 0.001 | 0.874 | 0.931 | -0.004 | 0.397 | 0.514 |
| parstriangularis (left) | 0.007 | 0.196 | 0.442 | 0.005 | 0.315 | 0.472 |
| pericalcarine (left) | 0.010 | 0.101 | 0.303 | 0.004 | 0.449 | 0.546 |
| postcentral (left) | -0.002 | 0.677 | 0.856 | -0.007 | 0.173 | 0.346 |
| posteriorcingulate (left) | 0.003 | 0.635 | 0.824 | -0.003 | 0.552 | 0.638 |
| precentral (left) | 0.004 | 0.460 | 0.743 | -0.008 | 0.085 | 0.181 |
| precuneus (left) | 0.005 | 0.274 | 0.533 | -0.004 | 0.331 | 0.472 |
| rostralanteriorcingulate (left) | -0.011 | 0.035 | 0.231 | -0.011 | 0.033 | 0.080 |
| rostralmiddlefrontal (left) | -0.007 | 0.114 | 0.312 | -0.005 | 0.247 | 0.398 |
| superiorfrontal (left) | -0.001 | 0.840 | 0.931 | -0.006 | 0.195 | 0.358 |
| superiorparietal (left) | 0.008 | 0.094 | 0.296 | -0.003 | 0.523 | 0.616 |
| superiortemporal (left) | -0.010 | 0.031 | 0.231 | -0.017 | < 0.001 | **0.002** |
| supramarginal (left) | 0.003 | 0.548 | 0.769 | -0.011 | 0.028 | 0.075 |
| frontalpole (left) | -0.001 | 0.867 | 0.931 | -0.001 | 0.812 | 0.839 |
| transversetemporal (left) | 0.003 | 0.591 | 0.812 | -0.004 | 0.435 | 0.542 |
| insula (left) | 0.003 | 0.511 | 0.767 | -0.013 | 0.005 | **0.018** |
| bankssts (right) | -0.019 | 0.001 | **0.013** | -0.024 | < 0.001 | **< 0.001** |
| caudalanteriorcingulate (right) | -0.012 | 0.041 | 0.231 | -0.015 | 0.011 | **0.032** |
| caudalmiddlefrontal (right) | -0.010 | 0.069 | 0.253 | -0.011 | 0.039 | 0.092 |
| cuneus (right) | 0.010 | 0.064 | 0.252 | 0.008 | 0.142 | 0.292 |
| entorhinal (right) | -0.002 | 0.713 | 0.856 | -0.015 | 0.009 | **0.028** |
| fusiform (right) | -0.007 | 0.155 | 0.364 | -0.016 | < 0.001 | **0.003** |
| inferiorparietal (right) | -0.013 | 0.010 | 0.129 | -0.017 | < 0.001 | **0.003** |
| inferiortemporal (right) | -0.010 | 0.042 | 0.231 | -0.017 | < 0.001 | **0.003** |
| isthmuscingulate (right) | -0.007 | 0.208 | 0.442 | -0.005 | 0.304 | 0.472 |
| lateraloccipital (right) | 0.000 | 0.934 | 0.963 | -0.004 | 0.364 | 0.490 |
| lateralorbitofrontal (right) | 0.000 | 0.996 | 0.996 | -0.006 | 0.187 | 0.354 |
| lingual (right) | 0.009 | 0.134 | 0.355 | 0.007 | 0.237 | 0.390 |
| medialorbitofrontal (right) | 0.016 | 0.001 | **0.013** | 0.004 | 0.385 | 0.508 |
| middletemporal (right) | -0.009 | 0.049 | 0.249 | -0.015 | 0.001 | 0.004 |
| parahippocampal (right) | 0.006 | 0.345 | 0.633 | 0.000 | 0.948 | 0.948 |
| paracentral (right) | -0.007 | 0.201 | 0.442 | -0.022 | < 0.001 | **< 0.001** |
| parsopercularis (right) | 0.001 | 0.865 | 0.931 | -0.001 | 0.814 | 0.839 |
| parsorbitalis (right) | 0.003 | 0.612 | 0.824 | 0.001 | 0.866 | 0.879 |
| parstriangularis (right) | 0.000 | 0.964 | 0.979 | 0.002 | 0.713 | 0.784 |
| pericalcarine (right) | 0.004 | 0.541 | 0.769 | 0.007 | 0.230 | 0.390 |
| postcentral (right) | -0.005 | 0.322 | 0.608 | -0.011 | 0.033 | 0.080 |
| posteriorcingulate (right) | -0.004 | 0.496 | 0.767 | -0.010 | 0.055 | 0.126 |
| precentral (right) | 0.006 | 0.221 | 0.456 | -0.006 | 0.220 | 0.382 |
| precuneus (right) | 0.002 | 0.702 | 0.856 | -0.004 | 0.312 | 0.472 |
| rostralanteriorcingulate (right) | -0.016 | 0.004 | 0.058 | -0.023 | < 0.001 | **< 0.001** |
| rostralmiddlefrontal (right) | -0.008 | 0.060 | 0.252 | -0.006 | 0.211 | 0.377 |
| superiorfrontal (right) | -0.006 | 0.146 | 0.358 | -0.014 | 0.001 | **0.006** |
| superiorparietal (right) | 0.006 | 0.252 | 0.504 | -0.005 | 0.328 | 0.472 |
| superiortemporal (right) | -0.008 | 0.090 | 0.296 | -0.012 | 0.007 | 0.025 |
| supramarginal (right) | -0.004 | 0.462 | 0.743 | -0.015 | 0.002 | **0.009** |
| frontalpole (right) | -0.005 | 0.435 | 0.743 | -0.005 | 0.431 | 0.542 |
| transversetemporal (right) | -0.011 | 0.054 | 0.252 | -0.014 | 0.010 | 0.029 |
| insula (right) | -0.002 | 0.688 | 0.856 | -0.019 | < 0.001 | **< 0.001** |
