## Supplemental Material for "The ratio of left atrial and ventricular volume as new marker of atrial cardiopathy with relevance for stroke risk and cognitive dysfunction"

**Supplementary material for Paper: Evaluating left atrial to ventricular volume ratio as a novel marker of atrial cardiopathy. Links to stroke risk and cognitive function.**

### 1. Detailed Methods

Calculating LAVi and LA:LV ratio in the UK Biobank

LAVi was calculated by taking maximum left atrial volume and dividing it by body surface area. LA:LV ratio was calculated by taking maximum left atrial volume and dividing it by left ventricular end diastolic volume.

Identifying Participants with AF, heart failure, LV dilation and reduced EF in the UK Biobank

AF was identified from two sources, the automated diagnosis of AF on the electrocardiogram (ECG) performed during the imaging visit and the diagnosis of ICD code *I48 Atrial fibrillation or flutter* before the date of the imaging in the linked health records.

Patients with heart failure were identified through the diagnosis of ICD code *I50 heart failure* before the date of the imaging in the linked health records. Cut offs for reduced EF (male: < 49 %, female: < 52 %) and increased left ventricle end diastolic volume indexed to BSA (LVEDVi) (male: > 108 ml/m2, female: > 96 ml/m2) were based on previously reported norm values for cardiac resonance imaging.[37]

Definition of endpoints in the UK Biobank

For analyzing associations with of atrial indices with ischemic stroke or transient ischemic attack a composite outcome was defined using the earliest occurrence of "Date of ischemic stroke" and "Date first reported transient cerebral ischemic attacks and related syndromes" to indicate the event date of brain ischemia. The imaging visit date served as the start to follow-up. Survival analysis was performed treating ischemic stroke/TIA as primary endpoint and death as a competing event. Participants experiencing neither event by 01.01.2025 were censored at that date.

For analyzing associations with cognitive function, nine cognitive tests, performed as part of the UK Biobank assessment were analyzed. The cognitive tests were performed as part of a touchscreen questionnaire on the date of the imaging visit. Detailed description of these tests can be found elsewhere.[48,49] Standardized values (Z-Scores) were calculated for each of the test scores. Results of trail making tests and reaction time were multiplied with -1 so that higher scores correlate with better cognitive function. Then Z-Scores were averaged to four cognitive domains; executive function (Tower Rearranging Test, Trail Making Test Part B), processing speed (Reaction Time Test, Digit Symbol Substitution Test, Trail Making Test Part A), memory (Numeric Memory Test, Paired Associate Learning Test, Prospective Memory Test), and reasoning (Fluid Intelligence Test, Matrix Pattern Completion Test) as has been done in previous studies.[48] The Z-Scores of the four domains were then averaged to receive an overall score for cognitive function.

Ascertainment of covariates in the UK Biobank

For age, we used the age at imaging visit, sex was taken as recorded during the first visit. Prior diseases were identified from the linked medical health records and encoded as present, when the first diagnosis in the health records occurred before the imaging visit. Details as to which ICD codes were included for each prior disease can be found in Table S1. Body mass index (BMI) was measured during the imaging visit. Smoking status and alcohol consumption were ascertained from the touchscreen questionnaire. Smoking status was classified as never, previous or current while alcohol consumption was classified in six categories namely never, special occasions only, one to three times a month, once or twice a week, three or four times a week and daily or almost daily. Physical activity was ascertained from the IPAQ activity questionnaire and classified into the IPAQ activity groups low, moderate and high. Townsend index as recorded at baseline was included as measure of financial deprivation. Education status was ascertained from the touchscreen questionnaire.

Associations of cortical volumes with LAVi and LA:LV ratio

Brain MRI was performed for a subset of participants in the UK Biobank and processed according to a standard pipeline. Details can be found under [https://biobank.ctsu.ox.ac.uk/crystal/crystal/docs/brain_mri.pdf]. As part of the processing pipeline, Desikan-Killiany parcellation was performed and cortical volumes calculated using freesurfer and made available to researchers. Total intracranial volume was also calculated as part of the processing pipeline. We analyzed data from the first imaging visit.

#### Identifying Participants with reduced LV EF and LV dilation in the USZ stroke cohort

Cut offs for reduced LV EF (male: < 52 %, female: < 54 %) and increased LVEDVi (male: > 74 ml/m$^2$, female > 61 ml/m$^2$) were based on guidelines of the American Society of Cardiology.[32,33] These slightly differ from the cut offs in CMR imaging[38] as volume estimates in CMR differ from estimates in echocardiography.[37]

#### Adding MRproANP to models based on LA:LV ratio

MRproANP was also extracted from health records of patients if assessed during in-hospital stay of the stroke patient. We fitted a logistic regression for the association of age, sex, LA:LV ratio and the logarithm of MR proANP levels (log(MR proANP) with the probability of AF being identified as cause of ischemic stroke. We then assessed the performance of this model for predicting AF as stroke cause using 10-fold cross validation.

### 2. Supplementary Tables

| Disease | ICD codes |
|---|---|
| Hypertension | I10 Essential (primary) Hypertension, I11 Hypertensive heart disease, I12 Hypertensive renal disease, I13 Hypertensive heart and renal disease, I15 Secondary hypertension |
| Diabetes | E10 Insulin dependent diabetes mellitus, E11 Non insulin dependent diabetes mellitus, E12 Malnutrition related diabetes mellitus, E13 Other specified diabetes mellitus, E14 unspecified diabetes mellitus |
| Vascular disease | I25 chronic ischaemic heart disease, I70 atherosclerosis, I74 arterial embolism and thrombosis, Date of myocardial infarction (algorithmically defined outcome) |
| Dyslipidemia | E78 Disorders of lipoprotein metabolism and other lipidaemias. |

**Table S1.** ICD codes used to define cardiovascular risk factors. Adapted from [50]

| Classification | Criteria |
|---|---|
| Atrial Fibrillation or Flutter | • Atrial fibrillation or flutter detected in 12 channel ECG during workup or documented in medical history prior to stroke<br>• No other plausible etiology for ischemic stroke identified |
| Other determined etiology | • Other cause of ischemic stroke identified (e.g. large vessel atherosclerosis, small vessel occlusion, vessel dissection etc.)<br>• A minimum of 48h of rhythm monitoring performed, without detection of atrial fibrillation or flutter.<br>• No atrial fibrillation or flutter documented in the medical history |
| Unknown etiology or incomplete workup | • Criteria for neither of the groups above met. |

**Table S2**. Criteria for classifying stroke etiology.

| | LAVi [ml/m²] | | | LA:LV ratio | | |
|---|---|---|---|---|---|---|
| | **Estimate** | **Std. Error** | **Pr(>\|t\|)** | **Estimate** | **Std. Error** | **Pr(>\|t\|)** |
| **(Intercept)** | 38.47 | 0.35 | < 0.001 | 0.5169 | 0.0040 | **< 0.001** |
| **Age (years)** | -0.67 | 0.07 | **< 0.001** | 0.0053 | 0.0007 | **< 0.001** |
| **Sex (male)** | -0.67 | 0.13 | **< 0.001** | -0.0683 | 0.0014 | **< 0.001** |
| **Hypertension** | 1.42 | 0.15 | **< 0.001** | 0.0122 | 0.0017 | **< 0.001** |
| **Diabetes** | -1.48 | 0.30 | **< 0.001** | 0.0051 | 0.0034 | 0.14 |
| **Vascular disease** | 1.99 | 0.35 | **< 0.001** | 0.0156 | 0.0040 | **< 0.001** |
| **Dyslipidemia** | -0.39 | 0.17 | **0.02** | 0.0037 | 0.0019 | 0.05 |
| **BMI (kg/m^2)** | 0.64 | 0.07 | **< 0.001** | 0.0166 | 0.0008 | **< 0.001** |
| **Smoking (Never)** | 0.89 | 0.34 | **0.009** | 0.0087 | 0.0039 | **0.03** |
| **Smoking (Previous)** | 0.88 | 0.35 | **0.01** | 0.0067 | 0.0040 | 0.09 |
| **Activity (low)** | -3.22 | 0.21 | **< 0.001** | -0.0100 | 0.0024 | **< 0.001** |
| **Activity (moderate)** | -1.92 | 0.13 | **< 0.001** | -0.0068 | 0.0015 | **< 0.001** |

**Table S3**. **Linear regression modeling LAVi and LA:LV ratio**. Age, and BMI were standardized. For Activity, high activity is the reference, for smoking, current smoking is reference. LAVi: left atrial volume indexed to body surface area, LA:LV left atrial to ventricle volume, BMI: Body mass index

| | LAVi | | | LA:LV ratio | | |
|---|---|---|---|---|---|---|
| | **Estimate** | **Std. Error** | **Pr(>\|t\|)** | **Estimate** | **Std. Error** | **Pr(>\|t\|)** |
| **(Intercept)** | -0.054 | 0.030 | 0.08 | 0.175 | 0.031 | < 0.001 |
| **Age (years)** | -0.058 | 0.006 | **< 0.001** | 0.041 | 0.006 | **< 0.001** |
| **Sex (male)** | -0.059 | 0.011 | **< 0.001** | -0.529 | 0.011 | **< 0.001** |
| **Hypertension** | 0.123 | 0.013 | **< 0.001** | 0.095 | 0.013 | **< 0.001** |
| **Diabetes** | -0.129 | 0.026 | **< 0.001** | 0.040 | 0.027 | 0.14 |
| **Vascular disease** | 0.173 | 0.031 | **< 0.001** | 0.120 | 0.031 | **< 0.001** |
| **Dyslipidemia** | -0.034 | 0.015 | **0.02** | 0.029 | 0.015 | 0.05 |
| **BMI (kg/m^2)** | 0.056 | 0.006 | **< 0.001** | 0.128 | 0.006 | **< 0.001** |
| **Smoking (Never)** | 0.077 | 0.030 | **0.009** | 0.068 | 0.030 | **0.03** |
| **Smoking (Previous)** | 0.077 | 0.030 | **0.01** | 0.052 | 0.031 | 0.09 |
| **Activity (low)** | -0.280 | 0.018 | **< 0.001** | -0.078 | 0.018 | **< 0.001** |
| **Activity (moderate)** | -0.167 | 0.011 | **< 0.001** | -0.052 | 0.012 | **< 0.001** |

**Table S4**. **Linear regression modeling standardized LAVi and LA:LV ratio.** Standardization of LAVi and LA:LV ratio was performed to improve comparability of associations. Age and BMI were also standardized. For smoking, current smokers are the reference. For Activity, high activity is the reference. LAVi: left atrial volume indexed to body surface area, LA:LV left atrial to ventricle volume, BMI: Body mass index.

| Ischemic stroke/TIA | | | |
|---|---|---|---|
| | **Hazard ratio** | **95% CI** | **P-value** |
| LAVi | 1.10 | 0.96 - 1.26 | 0.17 |
| LA:LV ratio | 1.14 | 1.00 - 1.30 | **0.0498** |
| **Cognitive function** | | | |
| | **Coefficient** | **95% CI** | **P -value** |
| LAVi | -0.008 | (- 0.017) - (0.002) | 0.10 |
| LA:LV ratio | -0.048 | (- 0.032) - (- 0.057) | **< 0.001** |
| **Atrial fibrillation as ischemic stroke cause** | | | |
| | **Log Odds ratio** | **95% CI** | **P-value** |
| LAVi | 1.6 | 1.2 - 2.1 | **< 0.001** |
| LA:LV ratio | 2.1 | 1.5 - 2.6 | **< 0.001** |

**Table S5.** Unadjusted Associations of left atrial volume index (LAVi) and left atrium to left ventricular volume (LA:LV) ratio with sequelae of atrial cardiopathy.

**Ischemic stroke/TIA**: Hazard ratios (HR) from cause specific hazard models for ischemic stroke with death as competing event. HR > 1 indicate increased hazard rate of ischemic stroke/TIA. For LAVi and LA:LV ratio two separate models were calculated. 249 participants had an ischemic stroke/TIA as an endpoint (TIA: 88, ischemic stroke: 161), 606 death. Median time under observation was 6.8 years.

**Cognitive function**: Linear regression coefficients for cognitive function as independent variable. Coefficients > 0 indicate association of larger volumes with better cognitive function. For LAVi and LA:LV ratio two separate models were calculated.

**Atrial fibrillation as ischemic stroke cause**: Log odds ratio for AF as stroke cause as independent variable in multivariate logistic regression. Coefficients > 0 indicate association of larger volumes with AF as stoke cause. For LAVi and LA:LV ratio two separate models were calculated each adjusted for age and sex. 289 ischemic stroke patients included, 76 with atrial fibrillation as stroke cause.

LAVi and LA:LV ratio were standardized in all models, making effect sizes more comparable.

LAVi: left atrial volume indexed to body surface area, LA:LV ratio: left atrial to ventricular volume ratio.

| | Hazard ratio | 95% CI | P-value |
|---|---|---|---|
| LAVi | 1.02 | 0.94 - 1.11 | 0.67 |
| LA:LV ratio | 1.04 | 0.95 - 1.13 | 0.40 |

**Table S6**. **Hazard ratios (HR) of cause specific hazard models for death with participants censored at ischemic stroke/TIA as competing event.** HR > 1 indicate increased hazard rate of death. For LAVi and LA:LV ratio two separate models were calculated each adjusted for the described covariates. All the parameters were standardized, making effect sizes somewhat comparable. 249 had an ischemic stroke/TIA as an endpoint, 606 death. Median time under observation was 6.8 years. LAVi: left atrial volume indexed to body surface area, LA:LV ratio: left atrial to ventricular volume ratio.

| | Hazard ratio | 95% CI (lower) | 95% CI (upper) | P - value |
|---|---|---|---|---|
| LAVi | 1.11 | 0.97 | 1.27 | 0.15 |
| LA:LV ratio | 1.15 | 1.01 | 1.30 | **0.03** |

**Table S7**. **Hazard ratios (HR) of subdistribution hazard models for ischemic stroke/TIA with death as competing event.** HR > 1 indicate increased hazard rate of ischemic stroke/TIA. For LAVi and LA:LV ratio two separate models were calculated each adjusted for the described covariates. All the parameters were standardized, making effect sizes somewhat comparable. 249 had an ischemic stroke/TIA as an endpoint, 606 death. Median time under observation was 6.8 years. LAVi: left atrial volume indexed to body surface area, LA:LV ratio: left atrial to ventricular volume ratio.

| | Hazard ratio | 95% CI (lower) | 95% CI (upper) | P - value |
|---|---|---|---|---|
| LAVi | 1.02 | 0.94 | 1.11 | 0.67 |
| LA:LV ratio | 1.04 | 0.95 | 1.12 | 0.41 |

**Table S8**. **Hazard ratios (HR) of subdistribution hazard models for death with participants censored at ischemic stroke/TIA.** HR > 1 indicate increased hazard rate of death. For LAVi and LA:LV ratio two separate models were calculated each adjusted for the described covariates. All the parameters were standardized, making effect sizes somewhat comparable. 249 had an ischemic stroke/TIA as an endpoint, 606 death. Median time under observation was 6.8 years. LAVi: left atrial volume indexed to body surface area, LA:LV ratio: left atrial to ventricular volume ratio.

| | Female | | Male | |
|---|---|---|---|---|
| | **LA:LV ratio** | **LAVi** | **LA:LV ratio** | **LAVi** |
| ROC AUC | 0.896 (0.836 - 0.955) | 0.853 (0.777 - 0.929) | 0.793 (0.701 - 0.884) | 0.751 (0.658 - 0.844) |
| Optimal cut off point | 0.85 (0.69 - 0.96) | 36 ml/$m^2$ (34 - 41) | 0.71 (0.55 - 0.86) | 35 ml/$m^2$ (25 - 36) |
| | N = 124, cases = 46 | | N = 165, cases = 30 | |

**Table S9. Area under the receiver operating curve (ROC AUC) and optimal cut off point for identifying atrial fibrillation as stroke cause stratified by sex.** Optimal cut off point calculated with Youden Index. Standard error calculated with 1000 bootstrap samples.

| | Log odds ratio | Std. Error | Pr(>\|z\|) |
|---|---|---|---|
| **(Intercept)** | -1.48 | 0.34 | < 0.001 |
| **Age** | 0.30 | 0.26 | 0.24 |
| **Sex (male)** | -0.66 | 0.45 | 0.14 |
| **LA:LV ratio** | 1.21 | 0.33 | **< 0.001** |
| **log(MR pro ANP)** | 1.36 | 0.30 | **< 0.001** |

**Table S10. Logistic regression with probability of atrial fibrillation being detected as ischemic stroke cause as outcome.** Age, LA:LV ratio and log(MR proANP) were standardized to have mean 0 and standard deviation of 1. N = 224, cases = 57

### 3. Supplemental Figures

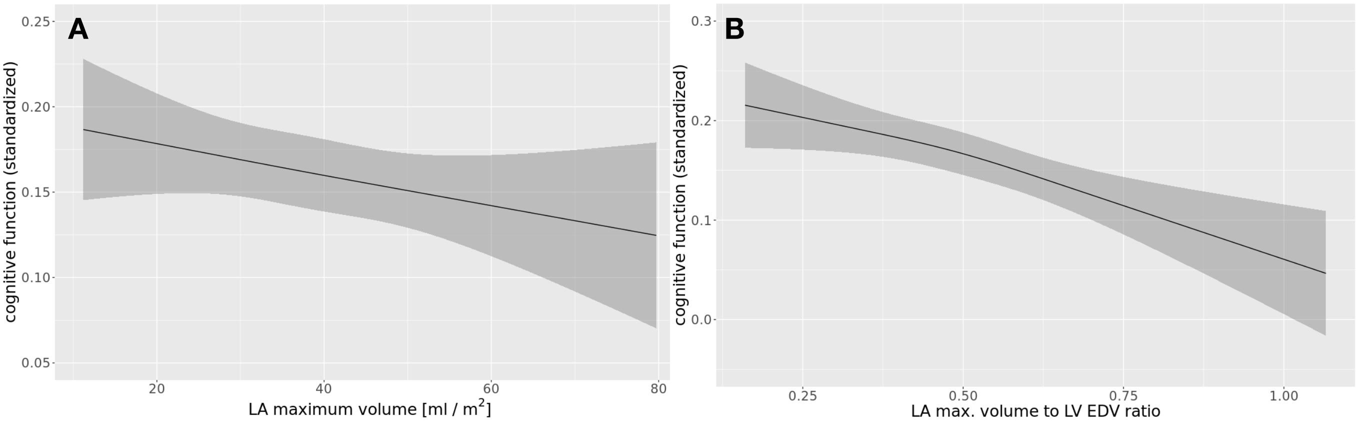


**Figure S1. Associations of left atrium volume indexed to body surface area (A) and Left atrium to left ventricle volume ratio (B) with cognitive function as modeled with restricted cubic splines with three nodes.** Models adjusted for covariates described in Table 1. For continuous covariates the parameter was fixed at the mean, for categorial variables the most common category was chosen. Neither of the parameters showed significant nonlinear components (A: p = 0.96, B: p = 0.51). LA maximum volume: left atrium maximum volume, LA max. volume to LV EDV ratio: left atrium maximum volume to left ventricle end diastolic volume ratio.

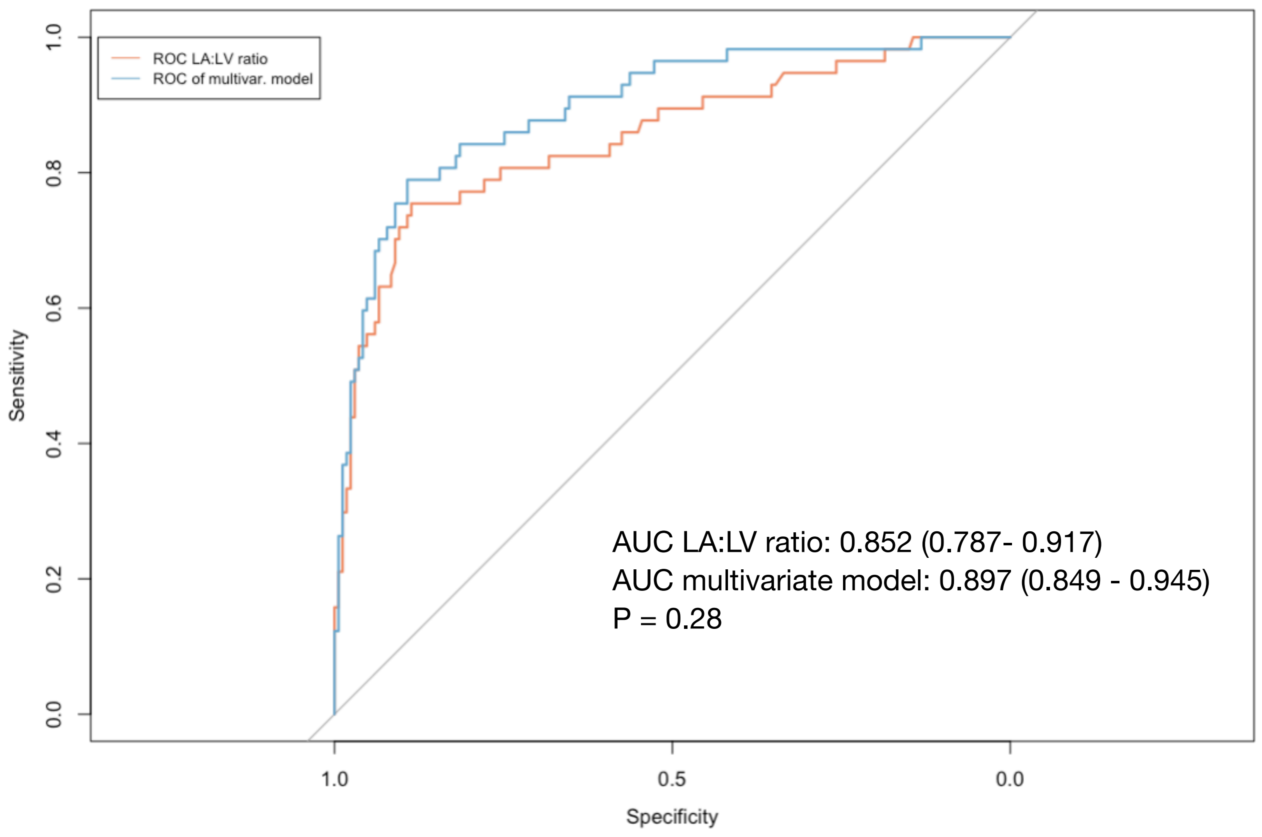


**Figure S2**. **Receiver operating characteristic curves for LA:LV ratio and multivariate model described in table S8 for detecting atrial fibrillation as stroke cause.** N = 224, cases = 57. Area under the curve (AUC) with 95 % CI is shown in the bottom right. Confidence intervals were calculated based on 1000 bootstrap samples. P value calculated with DeLongs's test. LAVi: Left atrial volume indexed to body surface area. LA:LV ratio: Left atrial to ventricle volume ratio, multivar.: multivariate, ROC: Receiver operating characteristic.
